# Understanding transfer of learning from an online self-directed learning and clinical decision support resource (BMJ Best Practice) for health professions education: a capability approach perspective

**DOI:** 10.1101/2021.07.18.21260229

**Authors:** Da Zhang, Li Xiao, Jingqi Duan, Xinxin Chang, Kieran Walsh, John Sandars, Jeremy Brown, Xiaorong Dang, Wei Shen, Junjie Du, Yanjie Cao

## Abstract

**Objective:** To understand transfer of learning of an online self-directed learning and clinical decision support resource for health professionals (BMJ Best Practice) informed by Sen’s capability approach.

**Design:** A semi-structured questionnaire was used to identify the extent to which BMJ Best Practice enabled participants to achieve their valued outcomes and the factors that enabled and constrained their achievement.

**Setting:** This study was carried out in a single centre, which is a tertiary care medical institution containing around 1500 inpatient beds.

**Participants:** 200 physicians at Air Force Medical Center were invited to take part in this study. 184 physicians completed the online questionnaire.

**Results:** 78 percent of physicians felt that BMJ Best Practice enabled them to achieve their valued outcomes and to apply their new knowledge to inform their practice. The main factors that constrained their achievement were technological.

**Conclusion:** Sen’s capability approach offers an innovative and useful model to further understand health professions education since it highlights the importance of the learner perspective of valued outcomes, including the difficulties associated with the effective transfer of learning in CPD.

**Strengths and limitations of this study:** This is a real-life study based on the experiences of physicians who are directly providing care to patients.

The study has strong theoretical foundations – being based as it is on Sen’s capability approach.

The study covers the vital subject of the application of medical knowledge in actual practice.

The research design is a cross-sectional survey; the study did not investigate whether physicians’ reflections on their valued outcomes and achievements might have changed over time.

This study was carried out in a single centre.

## INTRODUCTION

An essential aspect of all learning interventions is transfer – the extent to which new learning in one situation can be applied to another situation. Estimates from the world of management training indicate that transfer of learning to produce impact on performance can be as low as 30 – 40 percent^[1]^. The two most important factors that enable transfer to impact on performance appear to be whether the learning intervention (content and methods) meets the needs of the learner and the influence of the contextual environment within which the new learning is expected to be applied ^[2]^. There are similar concerns about the transfer of learning of continuing professional development (CPD) in health professions education ^[3, 4]^. An area of specific interest to our research team within the field of transfer is the increasing use of technology for CPD, especially where learning is self-directed^[5]^.

Sen’s capability approach has been increasingly recognized as a useful perspective for understanding and developing policy and practice in various educational contexts^[6]^.

We have recently developed a model for understanding transfer of learning that is based on Sen’s capability approach^[7]^. This model has a self-directed and learner-centered focus that considers how learners actively identify aspects of their daily practice that require further development through an educational experience. These areas for development are considered by the learners as being something that they value as being important (valued outcomes). The concept of value has recently been highlighted as having a more holistic and individual perspective on human development and learning than the use of the term learning needs ^[8]^.

Importantly from a capability approach perspective, understanding transfer of learning requires identification of the extent to which the learners’ valued outcomes can be achieved from both the educational experience and also the extent to which the learning through the experience can be applied to practice. In addition, it is important to understand the factors that enable and constrain the learner in achieving their valued outcomes.

BMJ Best Practice (http://bestpractice.bmj.com) is a unique self-directed online learning and clinical decision support resource to support diagnosis and treatment decisions, including shared decision-making with patients and their carers. Research evidence, guidelines and expert opinion are presented in a step-by-step approach. The resources are reviewed by experts to ensure relevance and quality of content. Transfer of the knowledge within this resource into actual clinical practice is essential to the implementation of evidence-based medicine.

We are not aware of previous research that has had a specific focus on understanding the complexity of transfer of learning in this area of online self-directed learning in health professions education, or the use of a capability approach perspective. The purpose of this research is to understand transfer of learning of an online self-directed learning and clinical decision support resource for health professionals informed by Sen’s capability approach.

## METHODS

### Participants and recruitment

Doctors and residents at Air Force Medical Center were invited to voluntarily take part in this study. Doctors were sent an introductory email by the lead researcher at Air Force Medical Center offering the opportunity of voluntarily participating in the research project. A Participant Information Sheet was attached to the email with further details of the study and how to register. Registration for the study will be done through completion of an Excel form, the data from which was used to email participants.

### Intervention

Participants were given access to BMJ Best Practice. To motivate participants to use the resources, participants received three reminders to use BMJ Best Practice during the study period.

### Evaluation

Since no previous questionnaire of transfer of learning informed by Sen’s capability approach was available, an online questionnaire was developed, with a mix of Likert type scales and open text questions to identify (a) the extent to which the learners’ valued outcomes can be achieved from the educational experience and how the learning through the experience can be applied to practice, and (b) the factors that enable and constrain the learner in achieving their valued outcomes. The questionnaire was pre-tested for its availability among 10 users in purpose of ensuring every participant could use it easily.

After 12 weeks of access to BMJ Best Practice, all participating doctors will be notified through Wechat (the most popular social media in China) with an invitation to complete the online questionnaire. Reminders to complete the survey were sent out to non-responders one and three weeks after the initial Wechat notification. Doctors were thanked for their participation and the study closed on the 17th week.

### Timeframe

Study registration was open for two weeks. The learning intervention period was 12 weeks and the questionnaire was administered after this.

### Data analysis

All questionnaires (completed and incompleted) were analyzed. Quantitative data from the questionnaires was exported and analysed using SPSS for descriptive frequencies and correlations between Likert scales of valued outcomes, achievement of new learning and application of new learning to practice.

Qualitative data from the questionnaires was exported to Excel and themes related to the factors in BMJ Best Practice that enable and constrain a learner to obtain and apply knowledge to meet their valued outcomes, and the types of barriers in the contextual environment for applying knowledge from BMJ Best Practice to inform their practice, was identified using Framework Analysis^[9]^.

### Data handling

Electronic data from the online questionnaire was stored on password protected computers. Hard copies of research project documentation were stored in a locked cabinet in a locked office. The procedures for handling, processing, storage, and destruction of data from the study were compliant with relevant national legislation.

### Ethical issues

All participants provided informed consent. This was obtained by providing a Participant Information Sheet and asking participants to indicate on the registration form that they give consent to take part. Participants needed to provide names, cellular phone numbers and email addresses to register with BMJ Best Practice and this personal identifying data was used to communicate with the participants for reminders and for the link to the online questionnaire. However, each participant was given a unique identifying number, and this was the only identifying data that was exported, analysed and stored. This process provided anonymity and confidentiality for information governance purposes. The study was approved by the Air Force Medical Center for Research Data (No. 2020-143-PJ01).

### Patient and public involvement statement

Patients and the public were not involved in this research.

## RESULTS

200 physicians at Air Force Medical Center were invited to take part in this study. 184 physicians including 20 interns, 78 residents, 64 attending doctors and 22 vice-directors and directors completed the online questionnaire. All the physicians had access to BMJ Best Practice.

### Evidence-based diagnosis

84.2% (155/184) of physicians had a valued outcome of being able to make an evidence-based diagnosis (Figure 1). 84.8% (156/184) of physicians had used BMJ Best Practice to help them make an evidence-based diagnosis. 71.1% of physicians “agreed” and 15.2% “strongly agreed” that the information provided by BMJ Best Practice had enabled them to achieve their valued outcome by reinforcing their existing knowledge of a topic. Also, (a) 66.3% of physicians “agreed” and 15.2% “strongly agreed” that the information provided by BMJ Best Practice provided them with new knowledge of a topic, (b) 71.2% of physicians “agreed” and 14.7% “strongly agreed” that the information provided by BMJ Best Practice on the topics was useful, and (c) 70.7% of physicians “agreed” and 14.7% “strongly agreed” that they could apply the information obtained from BMJ Best Practice to make an evidence-based diagnosis in clinical practice (Table 1).

**Table 1.** Extent of transfer of learning from BMJ Best Practice. (created by Da Zhang and permitted to reuse by all authors)

|  | Evidence-based<br>diagnosis | Evidence-based<br>treatment | Share<br>decision-making |
| --- | --- | --- | --- |
| The information provided by BMJ Best Practice helped reinforce your existing knowledge of the topic? | 4.02±0.58 | 4.06±0.55 | 3.95±0.63 |
| The information provided by BMJ Best Practice provided you with new knowledge of the topic? | 3.96±0.65 | 4.02±0.58 | 3.93±0.65 |
| The information provided by BMJ Best Practice on this topic was useful? | 4.00±0.60 | 3.99±0.60 | 3.99±0.58 |
| You could apply the information you obtained from BMJ Best Practice on this topic in your clinical practice? | 4.01±0.58 | 3.99±0.60 | 3.99±0.56 |

Likert Scale Response (5 = Strongly Agree, 4 = Somewhat Agree, 3 = Neutral, 2 = Somewhat Disagree, 1 = Strongly Disagree).

**Figure 1.**
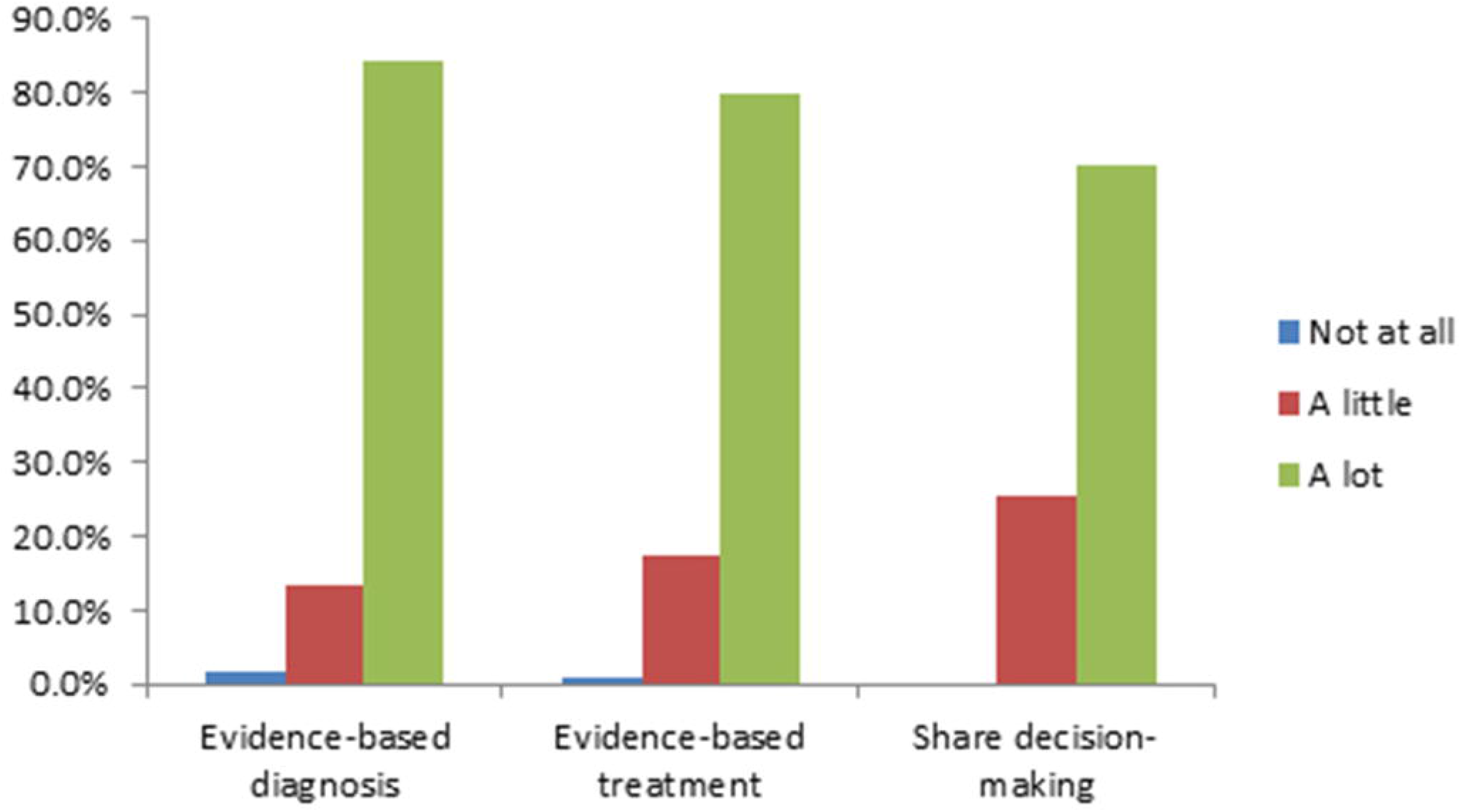
The extent to which users valued being able to: make an evidence-based diagnosis, start evidence-based treatment, and share decision-making. (created by Da Zhang and permitted to reuse by all authors)

16.3% (30/184) physicians encountered barriers that hindered them from using the information from BMJ Best Practice in making an evidence-based diagnosis. The most common barriers were technological barriers (such as slow speed), lack of local guidelines, and inability to find sufficiently detailed information about specific diseases.

There was no association between usage of BMJ Best Practice and the extent to which users value being able to make an evidence-based diagnosis (p>0.05). Usage of BMJ Best Practice was associated with users feeling that they have fewer barriers to making an evidenced based diagnosis (p<0.05). The more that users value being able to make an evidence-based diagnosis the more that they value BMJ Best Practice (p<0.05) (Table 2).

**Table 2.**
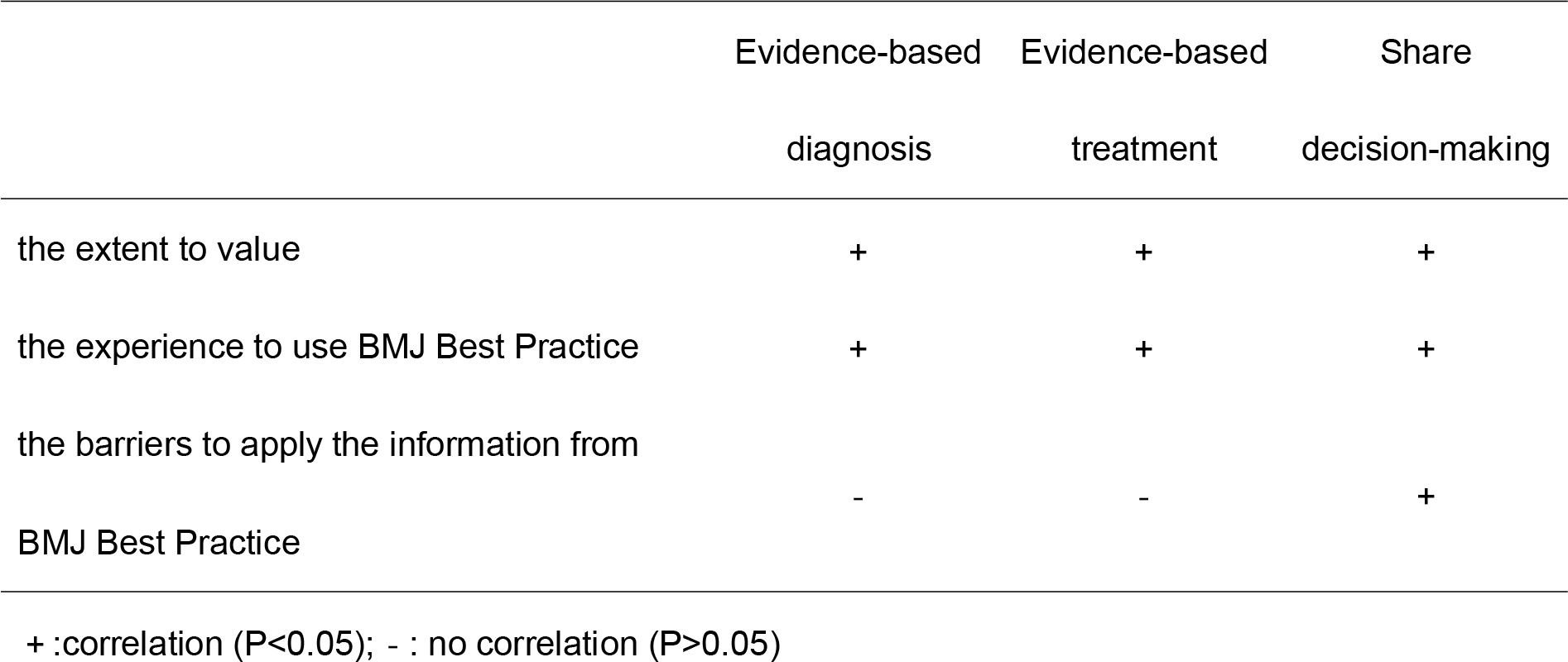
Factors in BMJ Best Practice that enable and constrain a learner to obtain and apply knowledge in clinical practice. (created by Da Zhang and permitted to reuse by all authors)

### Evidence-based treatment

79.9% (147/184) of physicians achieved a valued outcome of being able to provide evidence-based treatment (Figure 1). 80.4% (148/184) of physicians had used BMJ Best Practice to help them provide evidence-based treatment. 80.4% (148/184) physicians reported that BMJ Best Practice had enabled them to achieve their valued outcome of providing evidence-based treatment. 69.0% of physicians “agreed” and 16.8% “strongly agreed” that the information provided by BMJ Best Practice helped reinforce their existing knowledge. 65.8% of physicians “agreed” and 15.2% “strongly agreed” that the information provided by BMJ Best Practice provided them with new knowledge. 65.8% of physicians “agreed” and 15.2% “strongly agreed” that the information provided by BMJ Best Practice was useful. 66.3% of physicians “agreed” and 14.7% “strongly agreed” that they could apply the information obtained from BMJ Best Practice to provide evidence-based treatment in clinical practice (Table 1). 13.0% (24/184) of physicians encountered barriers that hindered them from using the information from BMJ Best Practice to provide evidence-based treatment. The barriers were similar to those related to making an evidence-based diagnosis. There was no association between usage of BMJ Best Practice and the extent to which users value being able to provide evidence-based treatment (p>0.05). Usage of BMJ Best Practice was not associated with users feeling that they have fewer barriers to providing evidenced based treatment (p>0.05). The more that users value being able to provide evidence-based treatment the more that they value BMJ Best Practice (p<0.05) (Table 2).

### Shared decision-making

70.1% (129/184) of physicians achieved a valued outcome of being able to share decision-making with patients and their carers (Figure 1). 65.8% (121/184) of physicians had used BMJ Best Practice to help them achieve their valued outcome of sharing decision-making with patients and their carers. 64.7% of physicians “agreed” and 14.1% “strongly agreed” that the information provided by BMJ Best Practice helped reinforce knowledge that would enable shared decision-making with patients and their carers. 64.1% of physicians “agreed” and 13.6% “strongly agreed” that the information provided by BMJ Best Practice provided them with new knowledge to share decision-making with patients and their carers. 65.2% of physicians “agreed” and 14.7% “strongly agreed” that the information provided by BMJ Best Practice was useful. 67.4% of physicians “agreed” and 14.1% “strongly agreed” that they could apply the information obtained from BMJ Best Practice to share decision-making (Table 1). 13.0% (24/184) of physicians encountered barriers that hindered them from using the information from BMJ Best Practice to share decision-making with patients and their carers. The barriers were similar to those related to making an evidence-based diagnosis.

There was an association between the extent to which users value shared decision making and the extent to which they use BMJ Best Practice (p<0.05). There was an association between lack of barriers to using BMJ Best Practice in making shared decisions and the extent to which users use BMJ Best Practice to make shared decisions (p<0.05) (Table 2).

## DISCUSSION

There has been increasing interest in transforming medical education to ensure that it is appropriate to meet the healthcare needs of the twenty-first century^[10, 11]^. We consider that Sen’s capability approach can provide an innovative and useful model to further understand health professions education since it highlights the importance of the learner perspective of valued outcomes, including the difficulties associated with the effective transfer of learning in CPD.

Diagnosis, treatment and sharing decision-making with patients and their carers are key processes in clinical practice. In this study, about 80 percent of physicians agreed that BMJ Best Practice enabled a learner to achieve their valued outcomes that were identified in daily practice and to apply their new knowledge to inform their practice. The extent to which physicians valued evidence-based medicine and their experience in using BMJ Best Practice were factors that enabled them to achieve their valued outcomes and apply their knowledge. Physicians who had not used BMJ Best Practice or did not value evidence-based medicine encountered more barriers in the contextual environment of clinical practice.

Our study design and data have several strengths. This is a real-life study based on the experiences of physicians who are directly providing care to patients. The study also has strong theoretical foundations – being based as it is on Sen’s capability approach. Lastly it covers the vital subject of the application of medical knowledge in actual practice – in the important domains of diagnosis, management, and shared decision making.

Our study design and data had a few limitations. The research design is a cross-sectional survey. The study did not investigate whether physicians’ reflections on their valued outcomes and achievements might have changed over time. This study was carried out in a single centre, which is a tertiary care medical institution containing around 1500 inpatient beds and about 400 internal medicine physicians. Lastly there were some technical problems in the use of BMJ Best Practice in this centre – which might have acted as an additional barrier to achieving valued outcomes and the transfer of knowledge.

In this research, the majority of physicians felt that BMJ Best Practice helped learners to achieve valued outcomes that were identified in daily practice and to apply their new knowledge in practice. Sen’s capability approach offers an innovative and useful model to further understand health professions education since it highlights the importance of the learner perspective of valued outcomes, including the difficulties associated with the effective transfer of learning in CPD.

## Data Availability

The data are stored locally, please contact the corresponding author to discuss.

## Acknowledgements

We would like to thank all the physicians and all our colleagues involved in the BMJ Best Practice Study, especially those participating in the study.

## Funding

This project was supported by the Military Logistics Special Project for Health Care (18BJZ07)

## Competing interests

KW works for BMJ which produces the resource BMJ Best Practice.

## Availability of data and materials

The datasets collected in this study are available from the corresponding author on reasonable request.

## Ethics approval and consent to participate

The study was approved by the Air Force Medical Center for Research Data (No. 2020-143-PJ01).

## Each author’s contribution

Da Zhang led the study; made substantial contributions to the acquisition, analysis, or interpretation of data for the work; wrote the first draft and revised it critically for important intellectual content; approved the final version; and agrees to be accountable for all aspects of the work.

Li Xiao helped conduct the study; made substantial contributions to the acquisition, analysis, or interpretation of data for the work; revised it critically for important intellectual content; approved the final version; and agrees to be accountable for all aspects of the work.

Jingqi Duan helped conduct the study; made substantial contributions to the acquisition, analysis, or interpretation of data for the work; revised it critically for important intellectual content; approved the final version; and agrees to be accountable for all aspects of the work.

Xinxin Chang helped conduct the study; made substantial contributions to the acquisition, analysis, or interpretation of data for the work; revised it critically for important intellectual content; approved the final version; and agrees to be accountable for all aspects of the work.

Kieran Walsh made substantial contributions to the conception or design of the work; revised it critically for important intellectual content; approved the final version; and agrees to be accountable for all aspects of the work.

John Sandars made substantial contributions to the conception or design of the work; revised it critically for important intellectual content; approved the final version; and agrees to be accountable for all aspects of the work.

Jeremy Brown made substantial contributions to the conception or design of the work; revised it critically for important intellectual content; approved the final version; and agrees to be accountable for all aspects of the work.

Xiaorong Dang made substantial contributions to the acquisition, analysis, or interpretation of data for the work; revised it critically for important intellectual content; approved the final version; and agrees to be accountable for all aspects of the work. Wei Shen made substantial contributions to the acquisition, analysis, or interpretation of data for the work; revised it critically for important intellectual content; approved the final version; and agrees to be accountable for all aspects of the work.

Junjie Du made substantial contributions to the acquisition, analysis, or interpretation of data for the work; revised it critically for important intellectual content; approved the final version; and agrees to be accountable for all aspects of the work.

Yanjie Cao contributed to the design; made substantial contributions to the acquisition, analysis, or interpretation of data for the work; revised it critically for important intellectual content; approved the final version; and agrees to be accountable for all aspects of the work.

## Open access

This is an open access article distributed in accordance with the Creative Commons Attribution Non Commercial (CC BY-NC 4.0) license, which permits others to distribute, remix, adapt, build upon this work non-commercially, and license their derivative works on different terms, provided the original work is properly cited, appropriate credit is given, any changes made indicated, and the use is non-commercial. See: http://creativecommons.org/licenses/by-nc/4.0/.

## Notes

### Author Declarations

The study was approved by the Air Force Medical Center for Research Data (No. 2020-143-PJ01).

## References

[1] Blume B D, Ford J K, Baldwin T T, et al. Transfer of Training: A Meta-Analytic Review[J]. Journal of Management, 2010,36(4):1065–1105.

[2] Teunissen P W, Watling C J, Schrewe B, et al. Contextual Competence: How residents develop competent performance in new settings[J]. Med Educ, 2021.

[3] Walsh K, San Da Rs J, Kapoor S S, et al. Getting NICE guidelines into practice: can e-learning help?[J]. Clinical Governance An International Journal, 2010,15(1):6–11.

[4] Wilcock P M, Janes G, Chambers A. Health care improvement and continuing interprofessional education: continuing interprofessional development to improve patient outcomes[J]. J Contin Educ Health Prof, 2009,29(2):84–90.

[5] Sandars J. Cost-effective e-learning in medical education[M]. Cost Effectiveness in Medical Education, 2021.

[6] Chiappero-Martinetti E, Osmani S, Qizilbash M, editors. The Cambridge Handbook of the Capability Approach. Cambridge University Press; 2020

[7] Sandars J, Sarojini H C. The capability approach for medical education: AMEE Guide No. 97[J]. Med Teach, 2015,37(6):510–520.

[8] Sandars J, Walsh K. The value of health professions education: the importance of understanding the learner perspective. Education for Primary Care. 2016 Jul 3;27(4):254–7.

[9] Legard R, Keegan J, Ward K. Qualitative research practice: a guide for social science students and researchers[J]. 2003.

[10] Bhutta Z A, Chen L, Cohen J, et al. Education of health professionals for the 21st century: a global independent Commission[J]. Lancet, 2010,375(9721):1137–1138.

[11] Frank J R, Snell L S, Cate O T, et al. Competency-based medical education: theory to practice.[J]. Medical Teacher, 2010,8(32):638–645.

